# Marker genes of incident type 1 diabetes in peripheral blood mononuclear cells of children: A machine learning strategy for large-p, small-n scenarios

**DOI:** 10.1101/2022.02.07.22270652

**Authors:** Kushan De Silva, Ryan T. Demmer, Daniel Jönsson, Aya Mousa, Andrew Forbes, Joanne Enticott

## Abstract

**Background and objective:** Type 1 diabetes (TID) is a complex, polygenic disorder, the etiology of which is not fully elucidated. Machine learning (ML) genomics could provide novel insights on disease dynamics while high-dimensionality remains a challenge. This study aimed to identify marker genes of incident T1D in peripheral blood mononuclear cells (PBMC) of children via a ML strategy attuned to high-dimensionality.

**Methods:** Using samples from 105 children (81 with incident T1D and 24 healthy controls), we analyzed microarray transcriptomics via a workflow consisting of three sequential steps: application of dimension reduction strategies on the processed transcriptome; ML on the reduced gene expression matrix; and downstream network analyses to demarcate seed nodes (statistically significant genes) and hub genes. Sixteen dimension-reduction algorithms belonging to three groups (3 tailored; 3 regularizations; 10 classic) were applied. Four ML algorithms (multivariate adaptive regression splines, adaptive boosting, random forests, XGB-DART) were trained on the reduced feature set and internally-validated using repeated, 10-fold cross-validation. Marker genes were determined via variable importance metrics. Seed nodes were identified by the ‘*OmicsNet*’ platform while nodes having above average betweenness, closeness, and degree in the network were demarcated as hub genes.

**Results:** The processed gene expression matrix comprised 13515 genes which was reduced to contain 1003 genes collectively selected by dimension reduction algorithms. All four ML algorithms on this reduced feature set attained perfect and uniform predictive performance on internal validation. On removal of redundancies, variable importance metrics identified 30 marker genes of incident T1D in this cohort, while Early Growth Response 2 (EGR2) was uniformly selected by all four ML algorithms as the most important marker gene. Network analyses classified all 30 marker genes as seed nodes. Additionally, we identified 14 hub genes, 7 of which were found to be marker genes of incident T1D elucidated by ML.

**Conclusions:** We identified marker genes of incident T1D in PBMC of children via a ML analytic strategy attuned to the high dimensional structure of microarrays, with downstream analyses providing high biological plausibility. The demonstrated ML strategy would be useful in analyzing other high-dimensional biomedical data for biomarker discovery.

## 1. INTRODUCTION

Type 1 diabetes (T1D) is a chronic, polygenic disorder with a multifactorial aetiology encompassing strong genetic, autoimmune and environmental components, although its natural history is not yet fully elucidated [1]. It accounts for nearly 10% of the global diabetes prevalence and is the most common form of diabetes among children [2]. Insulin secretory dysfunction and hyperglycemia being hallmarks of the disease, people with T1D require lifelong insulin therapy. Increased mortality rates and complications [3], concomitantly with reduced quality of life [4], frequently associate with T1D.

Pathogenesis of T1D is driven by T-cell mediated destruction of β-cells, whilst autoantibodies targeting insulin, glutamic acid decarboxylase 65 (GAD65), tyrosine phosphatase-related islet antigen 2 (IA-2) and zinc transporter-8 (ZNT8) tend to appear long before symptomatic onset of the disease, and are thus considered early biomarkers of T1D [5]. Current evidence suggests an increased susceptibility of individuals with human leukocyte antigen (HLA)-DR and HLA-DQ genotypes to T1D [6], while genome-wide association studies have discovered a large number of non-HLA, T1D-associated genes as well [7]. Moreover, T1D-discordant monozygotic twin studies have reinforced the impact of environmental triggers, ascribing a potentially causal role for epigenetic changes such as hypomethylated gene promoter regions, in T1D pathogenesis [8, 9]. While important advances have been made with respect to T1D along its continuum of care, further concerted efforts are required to identify biomarkers and facilitate early detection and management to prevent complications.

Precision medicine initiatives combining analytic approaches such as artificial intelligence (AI) with multi-omics data, have shown promise for yielding novel and valuable findings on diseases including diabetes [10, 11]. With rapid and continuous progress in the disciplines of big data, omics, and AI, multidisciplinary studies amalgamating these domains to address knowledge gaps pertaining to the genetic basis of complex diseases such as T1D, are now a reality. As the classical statistical paradigm is constrained in its capability to handle the intricacies associated with high-dimensional data, machine learning (ML) offers a viable alternative for analyzing omics data.

The large-p, small-n structure (p>>>n), also coined as the “curse of dimensionality”, in which the feature space (p) heavily outnumbers the observations (n), is inherent in omics data including in gene expression microarrays, microbiome compositional data, and single cell RNA-sequencing assays. Being counterfactual to the common phenomenon of large-n, small-p contexts, classical statistical models are challenged by this unique structure. Various techniques proposed to address the curse of dimensionality in omics data, are dominated by feature selection workflows [12] and regularization techniques [13], while de-novo algorithms tailored to omics data have also been developed [14, 15]. Although a single best method to deal with high-dimensionality does not exist, proposed approaches have unequivocally contributed to increasing the robustness of omics data analytics and biomarker discovery.

In a previous study, we demonstrated the viability of a strategy combining multiple feature selection algorithms with ML for elucidating the markers of prediabetes using an epidemiological cohort [16]. However, omics data such as gene expression microarrays present a formidable analytic challenge with an enormous feature space of a disproportionately smaller sample. Robustness of an analogous approach combining classic feature selection, regularization, and de-novo dimension reduction algorithms with ML upon omics data is hitherto untested.

Gene expression profiles of incident T1D could potentially consist of marker genes and reveal insights on causally-linked transcriptomic alterations associated with the onset of T1D. Identification of peripheral blood-based biomarkers of T1D can also be useful for clinical diagnostic and screening purposes. In a previous study, differential expression of interleukin 1 beta (IL1B), early growth response 3 (EGR3), and prostaglandin-endoperoxide synthase 2 (PTGS2) genes in peripheral blood mononuclear cells (PBMC) was associated with incident T1D [17]. The objective of the present study was to analyze the gene expression profile of PBMC with respect to incident T1D in children using a ML strategy amenable to large-p, small-n scenarios and to identify marker genes. Further, we determined the biological plausibility of the findings by conducting downstream analyses including constructing protein-protein interactions (PPI) networks and demarcating hub-and seed-nodes.

## 2. METHODS

The analytic workflow is illustrated in **S1 Figure**.

### 2.1. Data retrieval and processing

Data used in this study are available on the open-source National Center for Biotechnology Information Gene Expression Omnibus (NCBI GEO) [18, 19] platform with the unique GEO accession ID of GSE9006. Microarray data were retrieved via the ‘*getGEO*’ function of ‘*GEOquery*’ R package [20]. Of note, GSE9006 contains gene expression profiles of children with both T1D and T2D, measured on diagnosis at baseline and 4 months afterwards, following treatment. As per the objectives of this study and since the sample of children with T2D was also smaller, only T1D samples were included. This comprised 105 samples in total, of which 81 were children with T1D and 24 were healthy controls. Furthermore, as gene expression profiles change with treatment, only baseline data were eligible. Observations without corresponding gene symbol annotations were removed. In case of multiple probes hybridized to the same gene, their mean expression values were estimated across the samples to create a single row each for a given gene symbol. The gene expression matrix was transposed and three demographic features deemed important, namely, age (years), gender, and race (White/Other), were annotated to the transposed matrix. Gene expression values of this curated dataset was log_2_ Normalized to create symmetric distributions devoid of skewness (**S2 Figure**).

### 2.2. Dimension reduction

We applied 16 dimension-reduction algorithms belonging to three categories on the curated, transposed, and log_2_ Normalized gene expression matrix (**Table 1**). A brief description of these algorithms follows.

**Table 1:**
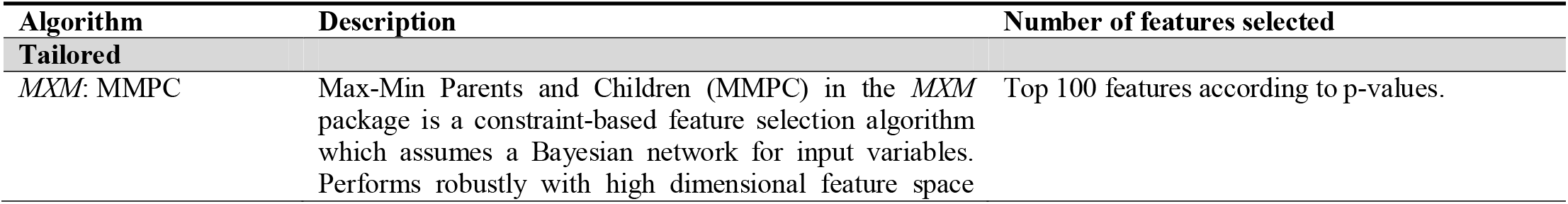
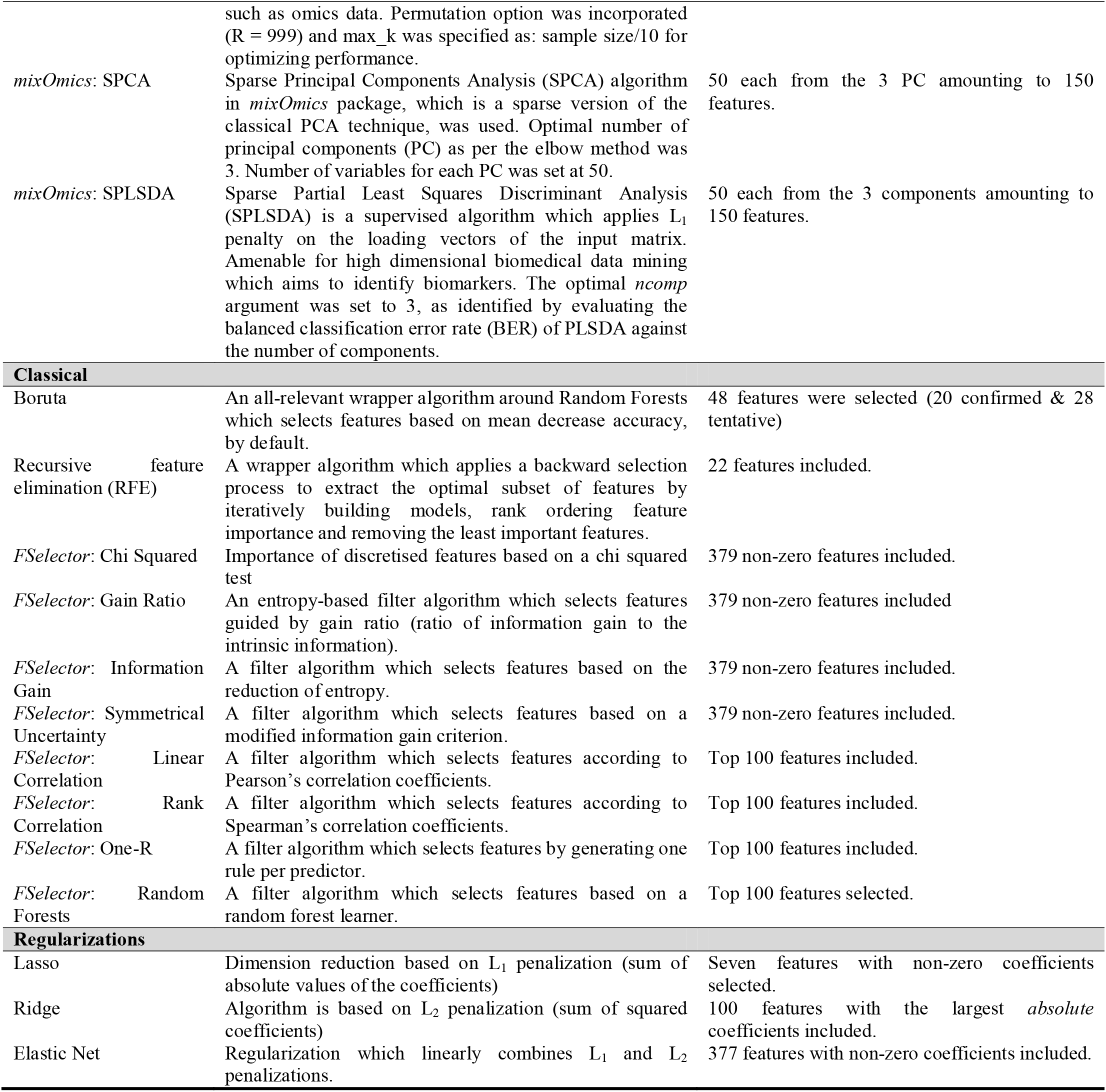
Summary of the 16 algorithms applied for dimension reduction and the outputs.

#### 2.2.1. De-novo algorithms tailored to handling the curse of dimensionality in omics (n = 3)

- *Sparse Principal Component Analysis (SPCA)*: Incorporated in the ‘*mixOmics*’ R-package [14], this algorithm selects features via singular value decomposition and lasso penalization on the loading vectors. The optimal number of principal components (PC) was determined via elbow method.
- *Sparse Partial Least Squares Discriminant Analysis (SPLSDA)*: Incorporated in the ‘*mixOmics*’ R-package [14], this supervised algorithm performs partial least squares-based feature reduction administering an L_1_ penalty on the loading vectors of the input matrix. Parameter tuning was performed to determine the optimal number of features and components by evaluating the balanced classification error rate (BER) of PLSDA against the number of features and components (**Figures 1-5**).
- *Max-Min Parents and Children (MMPC)*: Incorporated in the *MXM* R-package [15], this algorithm conducts a constraint-based feature selection, assuming a Bayesian network for input variables. The permutation option was activated (R = 999) and the max_k value was specified as: sample size/10 for optimizing the performance.

**Figure 1:**
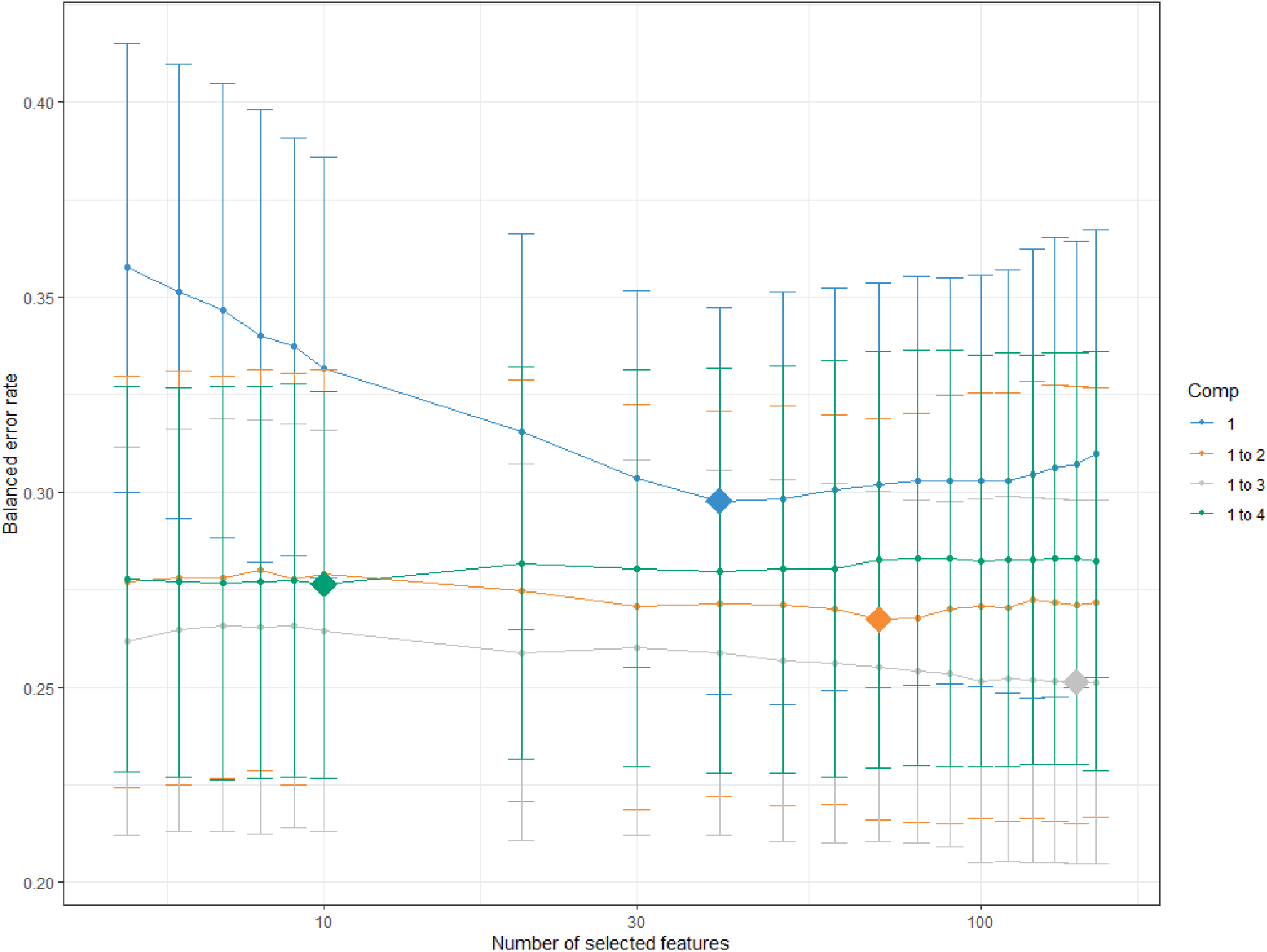
Parameter tuning to determine the optimal number of features and components for SPLSDA. Plot of the number of selected features against balanced error rate identified 150 features per component and ncomp = 3 as optimal.

**Figure 2:**
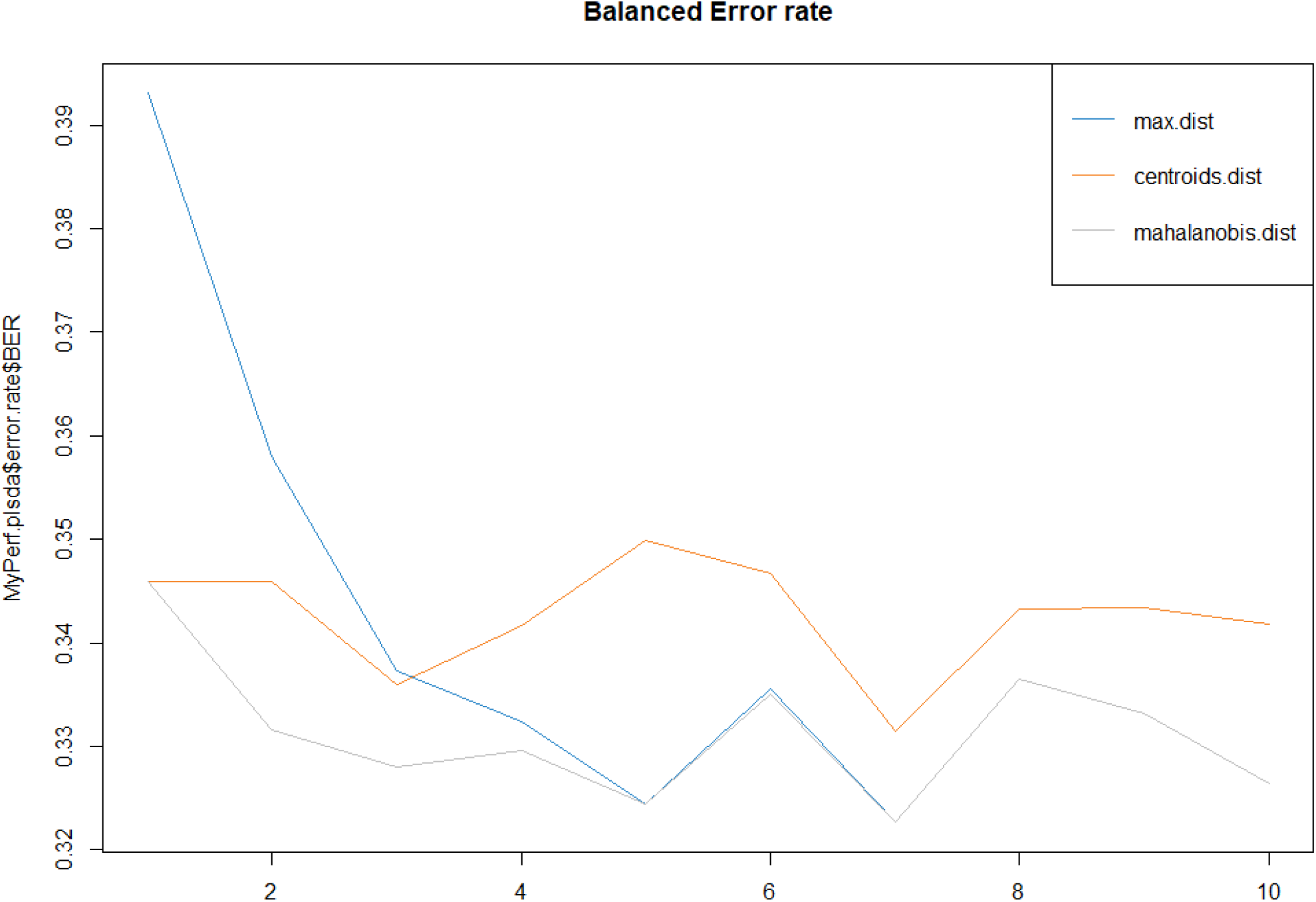
Parameter tuning to determine the optimal *ncomp* for SPLSDA: Plot of balanced classification error rate (BER) versus the number of components as per three prediction distances. The ncomp = 3 was selected as a reasonable criterion minimizing BER.

**Figure 3:**
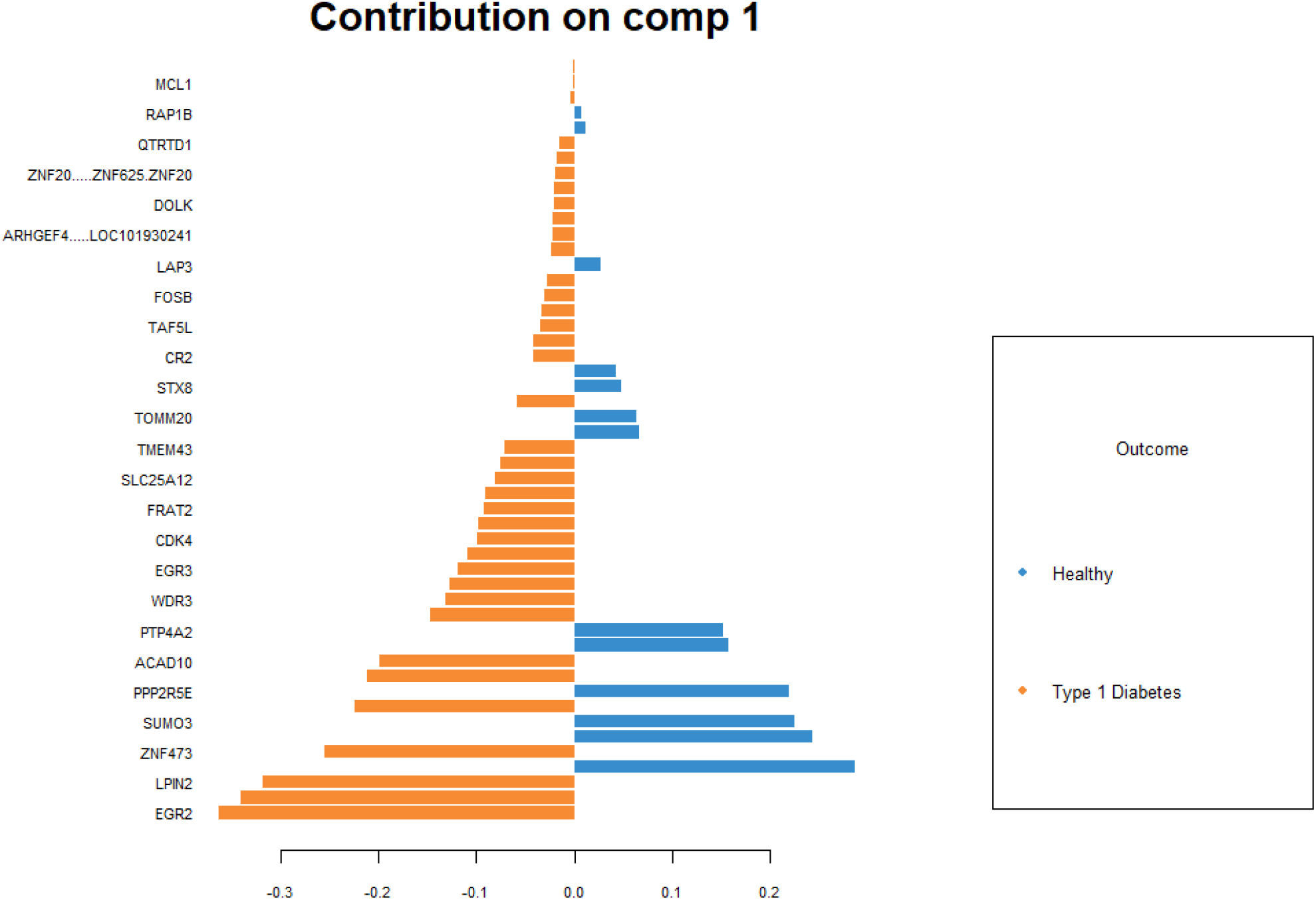
Features of the first component of SPLSDA output distributed between the two classes of the target variable.

**Figure 4:**
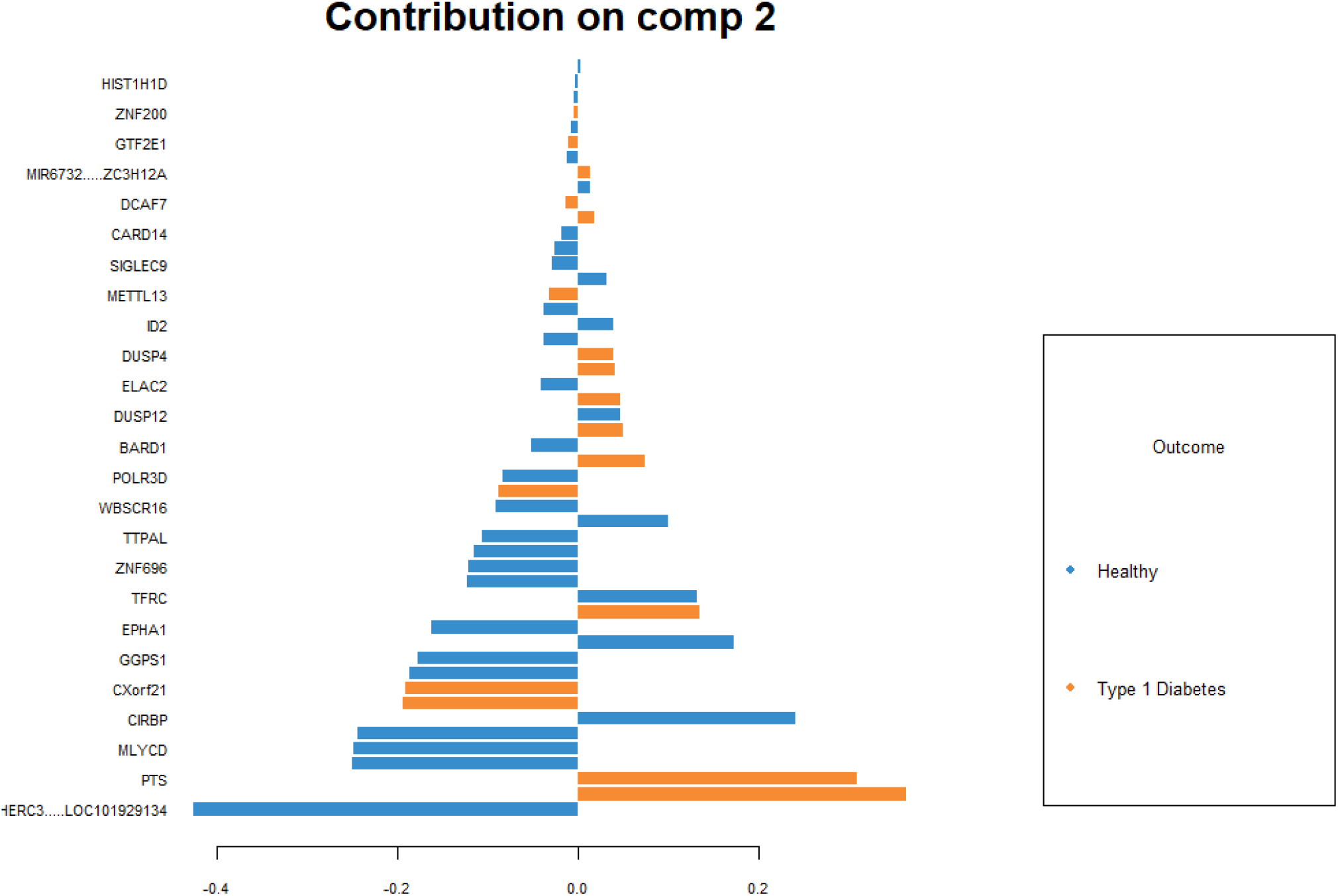
Features of the second component of SPLSDA output distributed between the two classes of the target variable.

**Figure 5:**
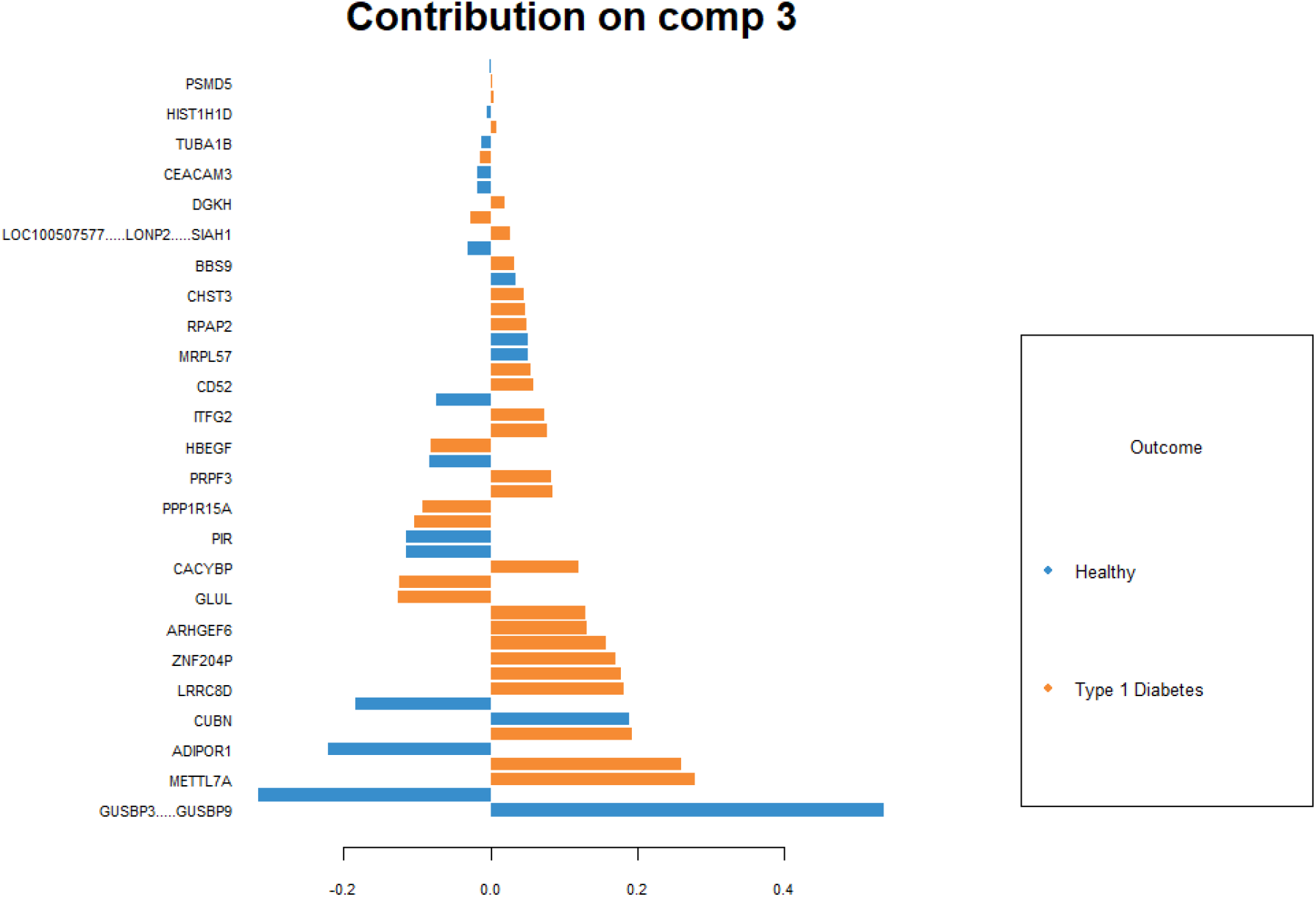
Features of the third component of SPLSDA output distributed between the two classes of the target variable.

#### 2.2.2. Regularization algorithms (n = 3)

- *Lasso*: This algorithm performs feature reduction based on L_1_ regularization, which adds a penalty term equal to the absolute value of the magnitude of coefficients.
- *Ridge*: This algorithm performs feature space reduction based on L_2_ penalization which adds a penalty term equal to the square of the magnitude of coefficients.
- *Elastic net*: This algorithm performs feature selection via a regularization method which linearly combines L_1_ and L_2_ penalties.

#### 2.2.3. Classic feature selection algorithms (n = 10)

- *Boruta*: This is an all-relevant wrapper algorithm around Random Forests which selects features based on mean decrease accuracy, by default [21].
- *Recursive feature elimination*: A greedy algorithm which extracts the best subset of features by iteratively building models, rank ordering feature importance and removing the least important features.
- *Information gain, gain ratio, and symmetrical uncertainty*: These entropy-based algorithms, incorporated in the *FSelector* R package [22], determine feature weights based on their correlation with the target variable.
- *Linear correlation and rank correlation*: These two correlation-based algorithms, integrated into the *FSelector* [22], utilise Pearson’s- and Spearman’s correlation coefficients, respectively, for feature selection.
- *Random forests*: A filter algorithm which selects features based on a random forest learner, found in *FSelector* [22].
- *One R*: A simple filter algorithm which selects features by generating one rule per predictor, found in *FSelector* [22].
- *Chi Squared*: This algorithm in *FSelector* [22] attributes importance to discretised features based on a chi squared test.

Features selected by each algorithm were compiled and repeated genes were removed. Demographic features (age, gender, race) were appended and the categorical variables (gender, race) were one-hot encoded to create dummy variables.

### 2.3. Machine learning

Machine learning with hyperparameter tuning was performed on the reduced feature set using four algorithms, namely, multivariate adaptive regression splines (MARS), adaptive boosting (AdaBoost), random forests (RF), and extreme gradient boosting-dropouts meet multiple additive regression trees (XGB-DART). The MARS algorithm is a non-parametric regression technique suited for high dimensional data, and an advancement of linear models, capable of automatic accounting for non-linearities and interactions within the feature space [23]. The AdaBoost is a meta-algorithm ensemble of decision trees which generates boosted classifiers by combining weak decision tree learners into a weighted sum [24]. The RF algorithm is an ensemble meta-learner based on bootstrap aggregating (“bagging”) of a multitude of decision trees accounting for overfitting [25]. The XGB-DART is a hybrid algorithm of two learners: XGBOOST is an ensemble algorithm [26] which collates a large number of decision trees with a small learning rate, while DART incorporates dropout techniques emanating from the deep neural network algorithmic paradigm in order to overcome overfitting [27]. For each algorithm, hyperparameter tuning was performed using *tuneLength* and *tuneGrid* commands of the *Caret* R package [28] and the metric for the optimal model selection was specified as “ROC”.

Internal validation of each model was performed by repeated, k-fold cross-validation (k = 10). The cross-validation approach has been the method of choice for internally validating omics-based ML models including gene expression analytics [29, 30]. In k-fold cross-validation, data are split into k non-overlapping folds. Each fold is set aside as test data while all other folds are combined into training sets. After a total of k models are fitted, performances are evaluated on the k test datasets and the mean performance is estimated. Repeated k-fold cross validation is an advancement which reduces the noise (error) of mean performance metrics attained by k-fold cross-validation.

The predictive performance of each model was evaluated using multiple performance metrics (**Table 2, S3 Figure**) and marker genes were identified via variable importance metrics (**Table 3, Figures 6-9**).

**Table 2:**
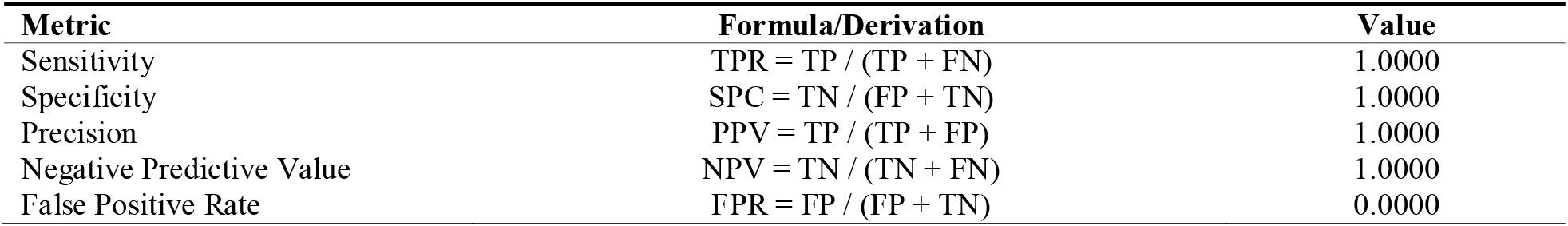
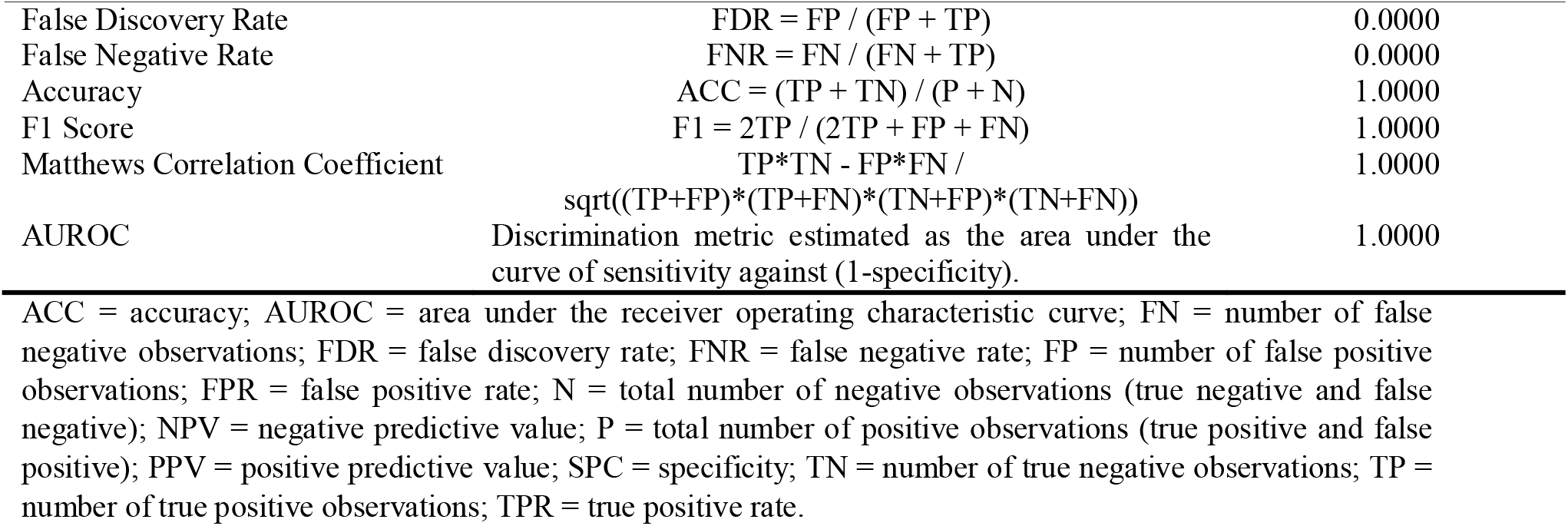
Predictive performance assessment metrics of internally-validated machine learning models. All four machine learning models achieved perfect performance on internal validation with uniform metrics given below.

**Table 3:**
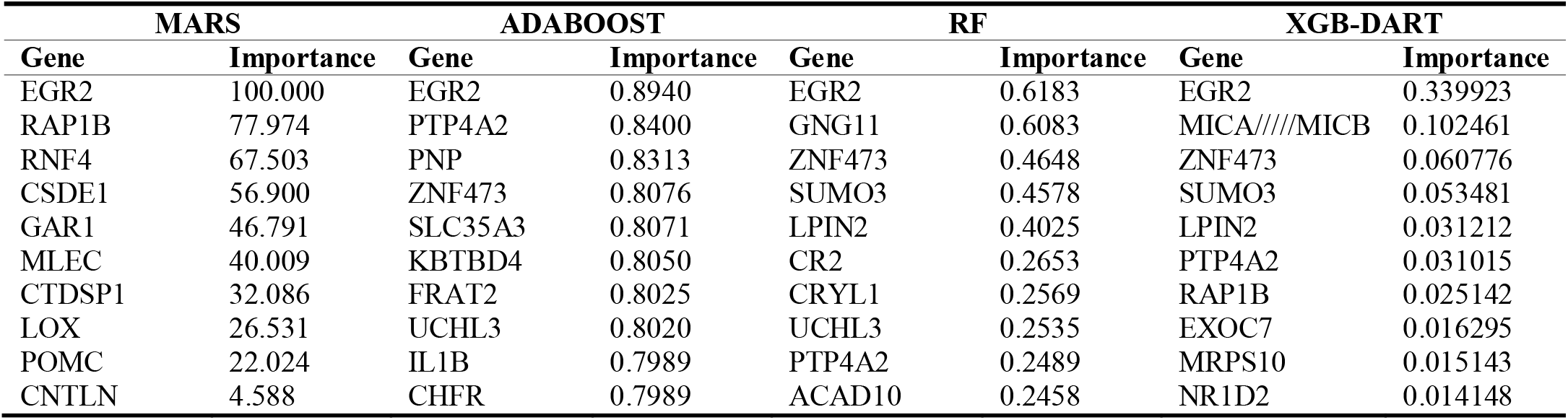
Variable importance metrics of machine learning models.

**Figure 6:**
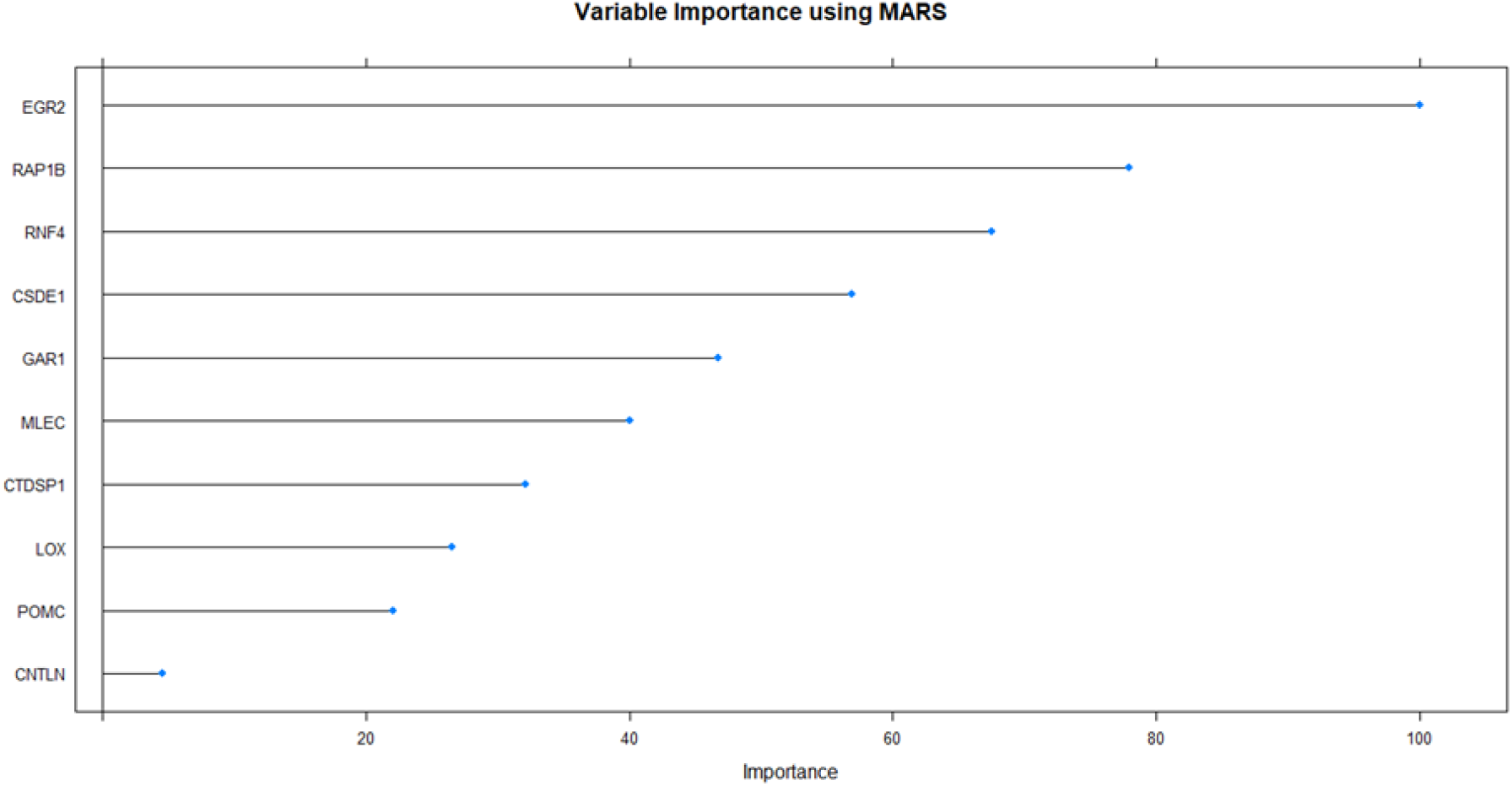
Variable importance plot of MARS model visualizing the top 10 most important features.

**Figure 7:**
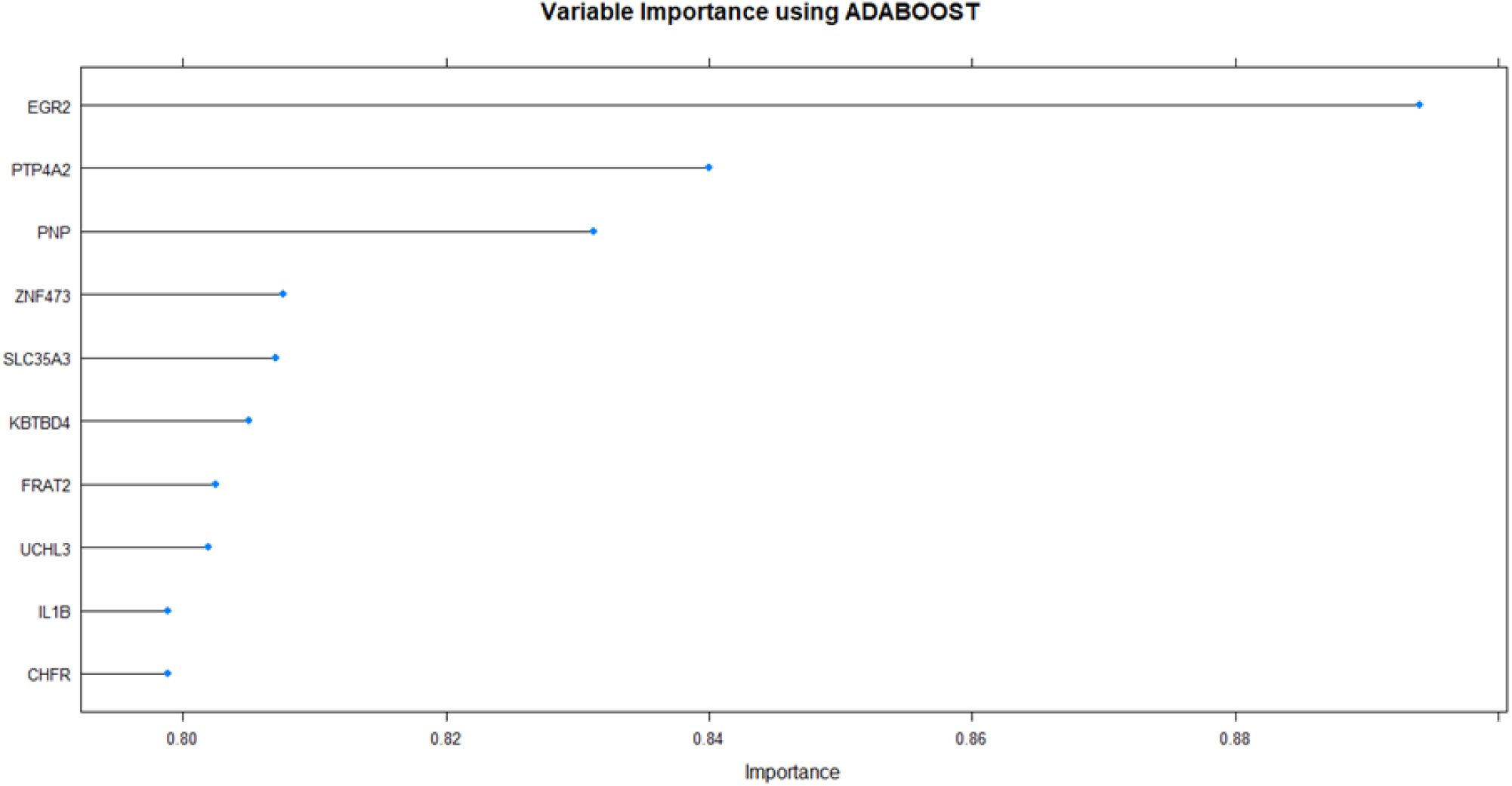
Variable importance plot of ADABOOST model visualizing the top 10 most important features.

**Figure 8:**
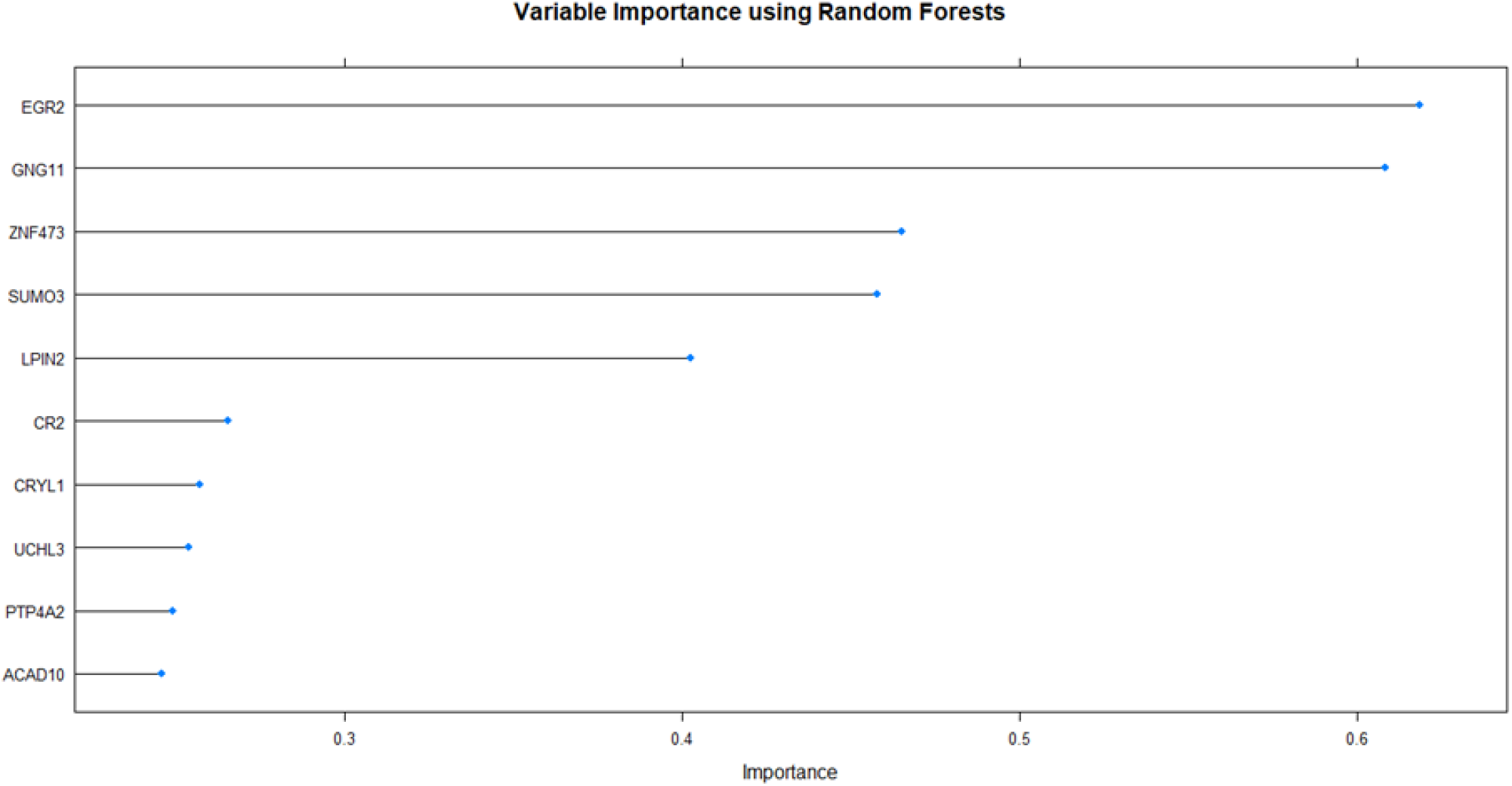
Variable importance plot of RF model visualizing the top 10 most important features.

**Figure 9:**
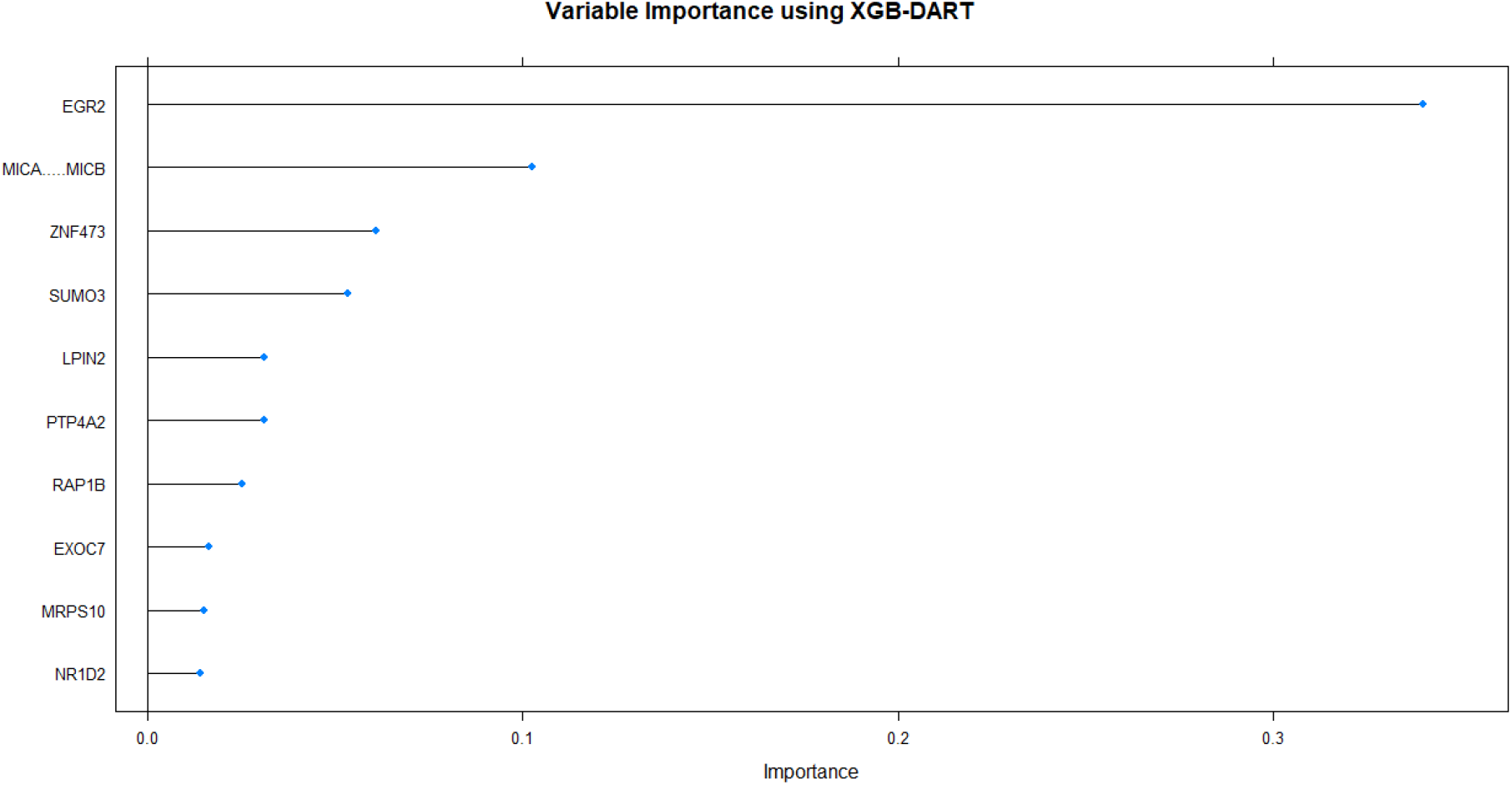
Variable importance plot of XGB-DART model visualizing the top 10 most important features.

### 2.4. Downstream analysis of marker genes

The ten most important genes identified by each ML model was compiled and repetitions were removed to generate the list of marker genes of incident T1D in PBMC of children. This list of genes was fed into *OmicsNet* [31, 32] which determines the significant genes (seeds) within the PPI network (**Figure 10**) by mapping input genes to the specified molecular interactions database.

**Figure 10:**
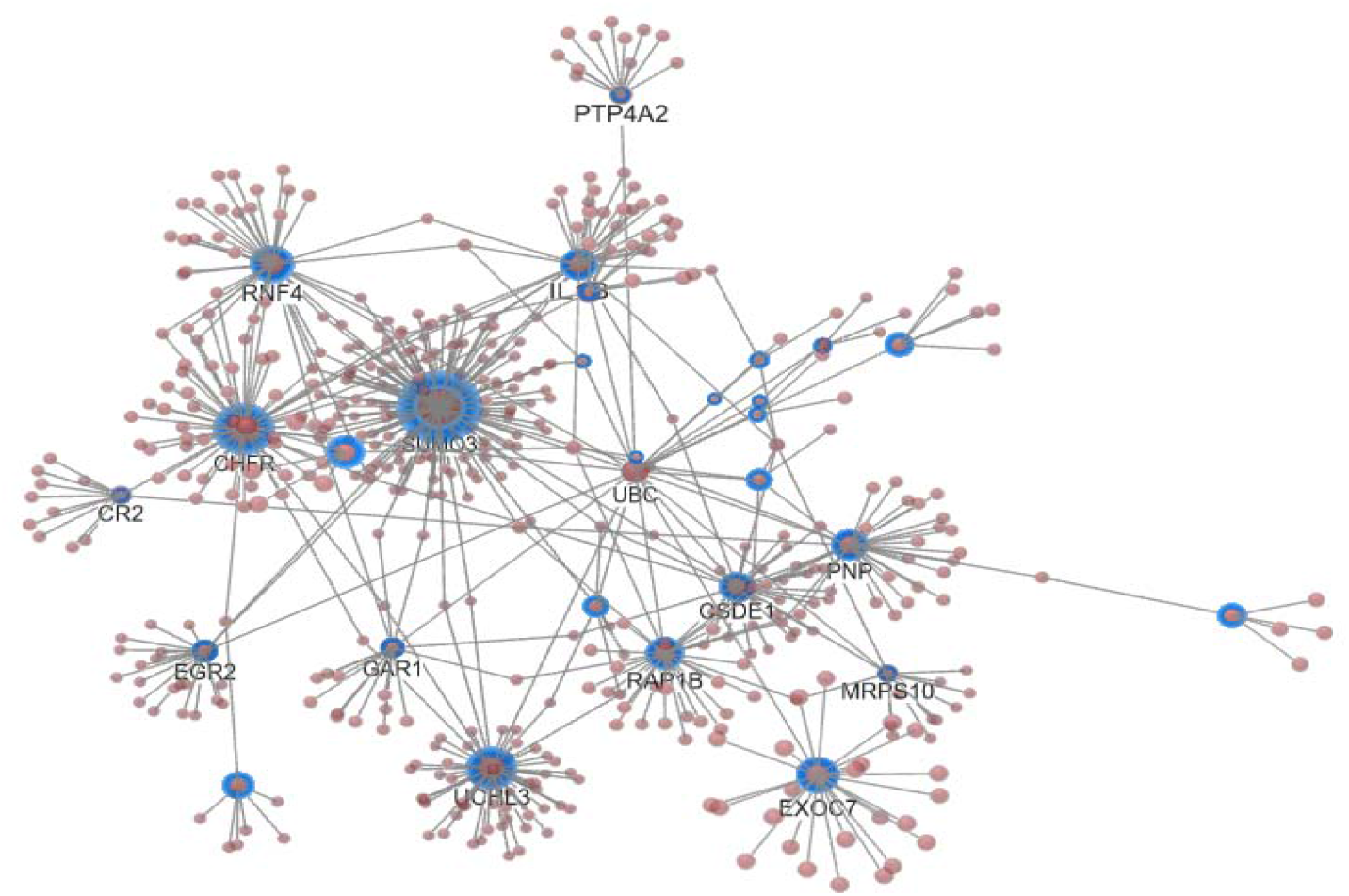
Protein-protein interactions network generated by *OmicsNet*. Seed nodes are highlighted (n = 30)

To demarcate the hub genes, we first created the gene-gene interactions network in GENEMANIA [33], which integrates an array of functional enrichments such as co-expression, pathways, physical interactions, co-localization, genetic interactions, and protein domain similarity into the network (**S4 Figure**). Next, the network was imported to *Cytoscape* [34] and its *Centiscape* plugin [35] was used to visualize the network and estimate topological metrics (**Figure 11**). As per Liu et al [36], nodes with above-average betweenness, closeness, and degree values were defined as hub genes (**Table 4**).

**Table 4:** Marker genes demarcated as hub nodes in the biological interaction network.

| Gene | Betweenness | Closeness | Degree |
| --- | --- | --- | --- |
|  | **mean = 70.938775510204** | **mean = 0.00869472439680752** | **mean = 7.83673469387755** |
| MLEC | 95.9111111111112 | 0.00961538461538461 | 9 |
| NR1D2 | 102.273835804718 | 0.01041666666666666 | 15 |
| CSDE1 | 131.437541053717 | 0.01041666666666666 | 17 |
| UCHL3 | 217.048665713371 | 0.0112359550561797 | 23 |
| RNF4 | 342.601101186394 | 0.0105263157894736 | 18 |
| RAP1B | 88.3532635664988 | 0.010204081632653 | 11 |
| CDV3* | 133.400057948587 | 0.010752688172043 | 11 |
| GTF2H3* | 96.9161763073529 | 0.010204081632653 | 10 |
| IMP3* | 151.888667704844 | 0.0103092783505154 | 16 |
| CTDSP2* | 124.136457333516 | 0.00961538461538461 | 11 |
| SP3* | 323.10581085581 | 0.0103092783505154 | 13 |
| H1-0* | 120.752832135185 | 0.01 | 10 |
| CUL2* | 255.441218748571 | 0.010752688172043 | 16 |
| PNP | 132.481177156177 | 0.00925925925925925 | 9 |

\*Predicted by GENEMANIA

**Figure 11:**
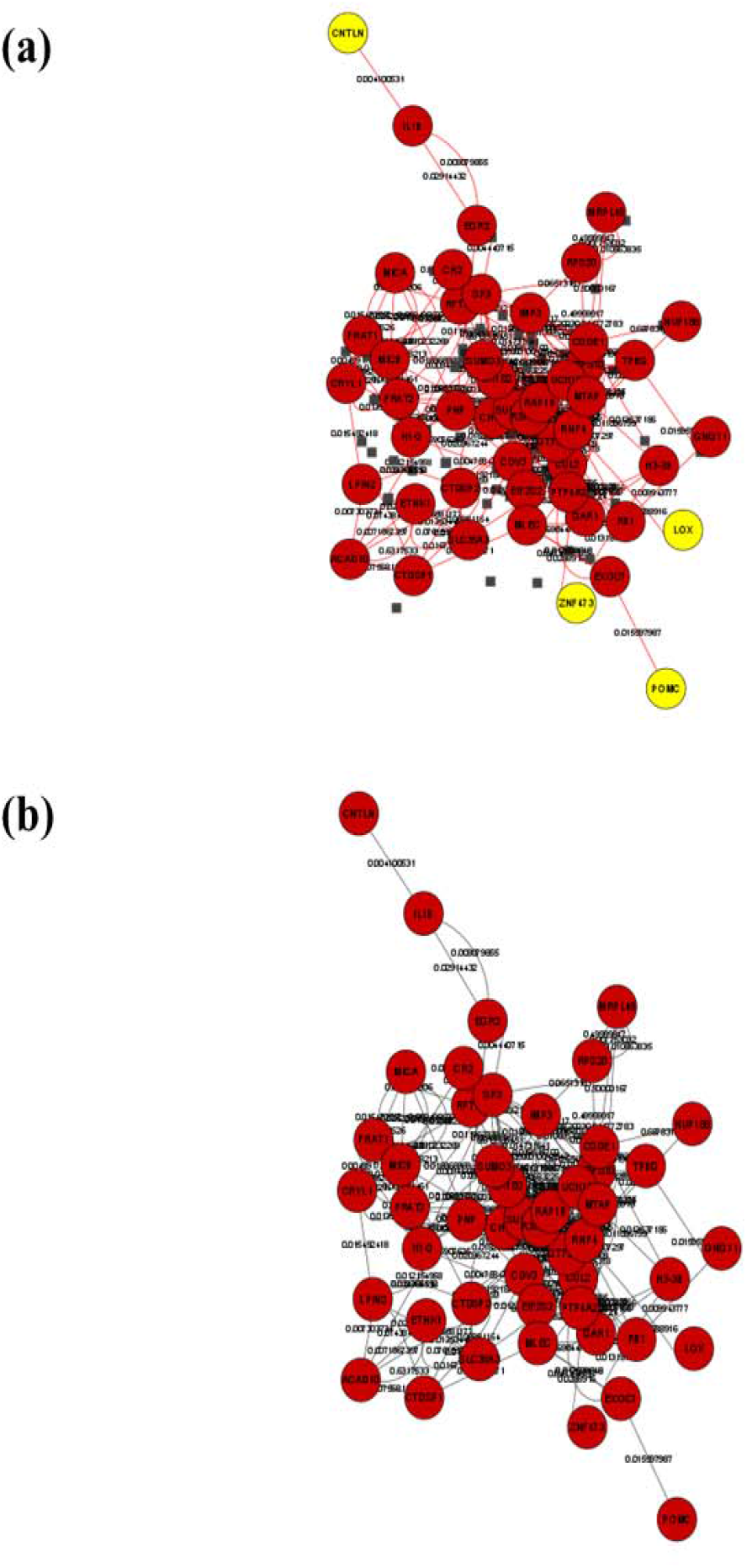
Two different presentations of the protein-protein interactions network visualized in *Cytoscape*.

## 3. RESULTS

The unprocessed gene expression matrix of PBMC at baseline consisted of 22283 probes measured on 105 children (81 with incident T1D and 24 healthy controls). After pre-processing, 13515 probes with unique gene symbols were retained (**S1 Table**). Age(years) distribution ranged from 2 – 17, with a mean (SD) of 9.81 (3.91) and a median (IQR) of 10.00 (6.00). Samples comprised 59 female and 46 male individuals. The dichotomized covariate of self-reported race was distributed as: 60 White and 45 Other (Black or African American = 8; selected more than one race = 5; other race = 3, race not reported = 29).

### 3.1. Features selected by tailored algorithms

Genes selected by running each of the 16 dimension-reduction algorithms are presented in **S2 Table**. For the SPCA algorithm, the optimal number of PC was found to be three according to the elbow method, and the 150 features selected (50 each from the 3 PC) are shown in **S2 Table**. The optimal number of features and components minimizing BER, with respect to the SPLSDA algorithm were found to be 150 and 3, respectively (**Figures 1 & 2**), and the selected 150 features (50 each from the 3 components) are illustrated in **Figures 3 - 5**. The **S3 Table** contains the complete feature ranking produced by the MMPC algorithm while the top 100 features ranked according to p-values that were selected for ML are shown in **S2 Table**.

### 3.2. Features selected by regularization algorithms

Seven candidate genes with non-zero coefficients (CSDE1, PPP2R5E, ZNF473, ACAD10, PNP, EGR2, LPIN2) were produced by Lasso regularization (**S4 Table**). The top 100 features with the largest *absolute* coefficients were selected from the ridge regularization algorithmic output (**S5 Table**). Elastic net algorithm converged with 377 non-zero feature coefficients (**S6 Table**).

### 3.3. Features selected by classic algorithms

Boruta algorithm selected 48 candidate genes (20 confirmed and 28 tentative) (**S7 Table**). Recursive feature elimination identified a sum of 22 variables maximizing cross-validated accuracy (**S2 Table**). Output produced by the eight algorithms incorporated in *FSelector* is given in **S8 Table**. Each of the three entropy-based algorithms (information gain, gain ratio, symmetrical uncertainty) converged with 379 non-zero coefficients. The top 100 features from the two correlation-based algorithms (rank- and linear-correlation) were selected for ML. We selected the top 100 features produced by random forests and One-R feature selection algorithms as well. Finally, the chi-squared algorithm generated 379 non-zero feature coefficients.

After removing redundancies, the collated feature set produced by all algorithms contained 1003 genes (**S9 Table**). The curated dataset containing these 1003 candidate genes and basic demographic variables (age, gender, race) used for ML is presented in **S10 Table**.

### 3.4. Machine learning

All four repeated, 10-fold cross-validated ML models converged attaining perfect and uniform predictive accuracy, classification-, and discrimination-metrics (**Table 2; S3 Figure**).

### 3.5. Variable importance

The 10 most important genes identified by the MARS model, in descending order, were: EGR2; RAP1B; RNF4; CSDE1; GAR1; MLEC; CTDSP1; LOX; POMC; CNTLN (**Figure 6**). The AdaBoost model elucidated: EGR2, PTP4A2, PNP, ZNF473, SLC35A3, KBTBD4, FRAT2, UCHL3, IL1B, CHFR, as the top 10 marker genes, in descending order (**Figure 7**). The RF model ranked: EGR2; GNG11; ZNF473; SUMO3; LPIN2; CR2; CRYL1; UCHL3; PTP4A2; ACAD10 genes as the top 10 marker genes, in diminishing importance (**Figure 8**). The most ten important genes identified by XGB-DART model, in descending order, were: EGR2; MICA/////MICB; ZNF473; SUMO3; LPIN2; PTP4A2; RAP1B; EXOC7; MRPS10; NR1D2 (**Figure 9**). Collation of the marker genes identified by the four ML models and removal of redundancies resulted in 30 candidate genes (**S11 Table**).

### 3.6. Network modules and seed nodes

Based on the 30 input genes, *OmicsNet* constructed a single subnetwork consisting of 504 nodes, 575 edges, and 23 modules (**Figure 10**), while all 30 marker genes were identified as seeds (statistically significant nodes of the PPI network) (**S12 Table**).

### 3.7. Hub genes

Details on the gene-gene interaction network constructed by GENEMANIA from the 30 input marker genes are provided in **S13 Table**. We identified 14 hub nodes within the PPI network which had above-average betweenness, closeness, and degree as per the topological values estimated by *Centiscape* (**S14 Table**). Seven of these hub nodes were found within the set of marker genes (MLEC, NR1D2, CSDE1, UCHL3, RNF4, RAP1B, PNP) while the remaining 7 hub genes were predicted by GENEMANIA (CDV3, GTF2H3, IMP3, CTDSP2, SP3, H1-0, CUL2).

## 4. DISCUSSION

In this study, we found a range of marker genes of incident T1D in PBMC of children, via an extensive ML workflow amenable for high dimensionality inherent in transcriptomics microarrays. Biological plausibility of our findings was confirmed by downstream analyses and appraisal of contemporary evidence. We envisage that the proposed ML strategy has the potential to integrate into various omics data types for an efficient biomarker discovery process.

### 4.1. Marker genes of incident T1D in PBMC of children: Biological plausibility

Noteworthily, the zinc finger transcription factor EGR2 was uniformly chosen by all four ML algorithms as the most important gene associated with incident T1D in this cohort. This is in tandem with previous studies in which EGR genes (both EGR2 and EGR3) were identified as highly perturbed at the onset of T1D [17]. There is emerging evidence on the role of EGR2 in the natural history of diabetes, through the development of insulin resistance which is increasingly observed in people with T1D [37] to the progression of complications [38, 39]. Recent studies which revealed crucial roles of EGR2 in the induction of T cell anergy and suppression of activities mediated by LAG3+ regulatory T cells (Tregs) are intriguing [40], given that Tregs are strongly implicated in T1D etiopathogenesis [41] and extensively investigated as therapeutic targets to ameliorate islet autoimmunity of T1D [42].

Our findings on the marker genes of incident T1D were congruent with previous studies which reported an innate inflammatory transcriptomic profile characterized by perturbations of genes such as CSDE1, EGR2, FRAT2, GNG11, IL1B, PTP4A2, SLC35A3, and UCHL3 [17, 43]. Moreover, there is topical evidence that RAP1B regulates glucose homeostasis [44], while the dysregulation of protein tyrosine phosphatases such as PTP4A2 (PRL2) is observed in several diabetes phenotypes [45]. Both MICA and MICB candidate genes belong to the HLA complex, which is collectively and consistently associated with T1D, contributing to 50% of its genetic risk [6]. An integrated analysis revealed that PNP is a strong diagnostic marker of T1D [46]. Furthermore, CR2 was found to be associated with abatacept-resistant, new-onset T1D, suggesting a likely role in B lymphocyte alterations [47]. Contemporary evidence also supports associations of POMC [48], and NR1D2 clock gene [49], with T1D.

### 4.2. Seeds and hub genes: Biological plausibility and implications

The inclusion of all 30 candidate genes as statistically significant nodes (seeds) in the PPI network was indicative of substantial gene-gene interactions between them. Further studies exploring these interactions could enhance our understanding of the genetic basis of T1D. Hub genes which appear as extensively and densely connected nodes in scale-free gene regulatory networks [50] tend to have important regulatory functions and offer value as potential therapeutic targets [51]. Therefore, hub genes identified in this analysis may be useful for guiding pharmacogenomics studies focused on T1D.

### 4.3. Methodological and analytic aspects

We observed a considerable degree of consistency with respect to the marker genes of T1D identified by ML, notwithstanding the essentially different dynamics between algorithms. However, some marker genes were uniquely identified by each algorithm, underscoring the need to train more than a single learner to gain robust and meaningful insights; a recommended practice in ML analytics [16]. It should also be noted that ML is intrinsically geared to optimizing prediction, although it is increasingly used to derive etiologic insights of diseases [52, 53]. Further studies focusing on the candidate genes identified in the current analysis, pragmatically combined with relatively larger samples are recommended, as they may provide novel insights on T1D pathogenesis. While the perfect predictive performance demonstrated by ML algorithms is encouraging, we underscore the caveats associated with the cross-validation approach, which is frequently used in omics analyses primarily owing to the large-p, small-n structure. Although we used repeated, k-fold cross-validation, which was found superior to other cross-validation techniques, they all have limitations [54]. Therefore, future research aiming to expand on this work should focus on externally validating our findings on different cohorts or internally validating on larger samples.

### 4.4. Applications

As high-dimensionality is frequently observed across different omics data typologies [55-57], we envision avenues for practical usage of the proposed ML strategy, beyond microarray transcriptomics.

## 5. CONCLUSIONS

In conclusion, we identified marker genes of incident T1D in PBMC of children via a ML analytic strategy attuned to the high dimensional structure of microarrays, with downstream analyses on seed nodes and hub genes providing high biological plausibility. The demonstrated ML strategy would have broader applications to studies aimed at biomarker discovery using high-dimensional, biomedical data.

## Data Availability

Data used in this study are freely available at the National Center for Biotechnology Information Gene Expression Omnibus (NCBI GEO) portal (https://www.ncbi.nlm.nih.gov/geo/), with the unique GEO accession ID of GSE9006.

## DECLARATIONS

### Ethics approval

Not required being a secondary analysis of publicly available, deidentified data.

### Author contributions

KDS performed data acquisition, pre-processing and curation of data, conducted the analyses and wrote this manuscript. KDS, RTD, AF and JE were responsible for study conceptualization and design, contributed to validating analyses, results and interpretation, and drafting the manuscript. DJ and AM contributed to study design, interpretation of data and critically and comprehensively revised the manuscript for important intellectual content.

