## Supplementary figures and images for "Marker genes of incident type 1 diabetes in peripheral blood mononuclear cells of children: A machine learning strategy for large-p, small-n scenarios"

### S1 Figure

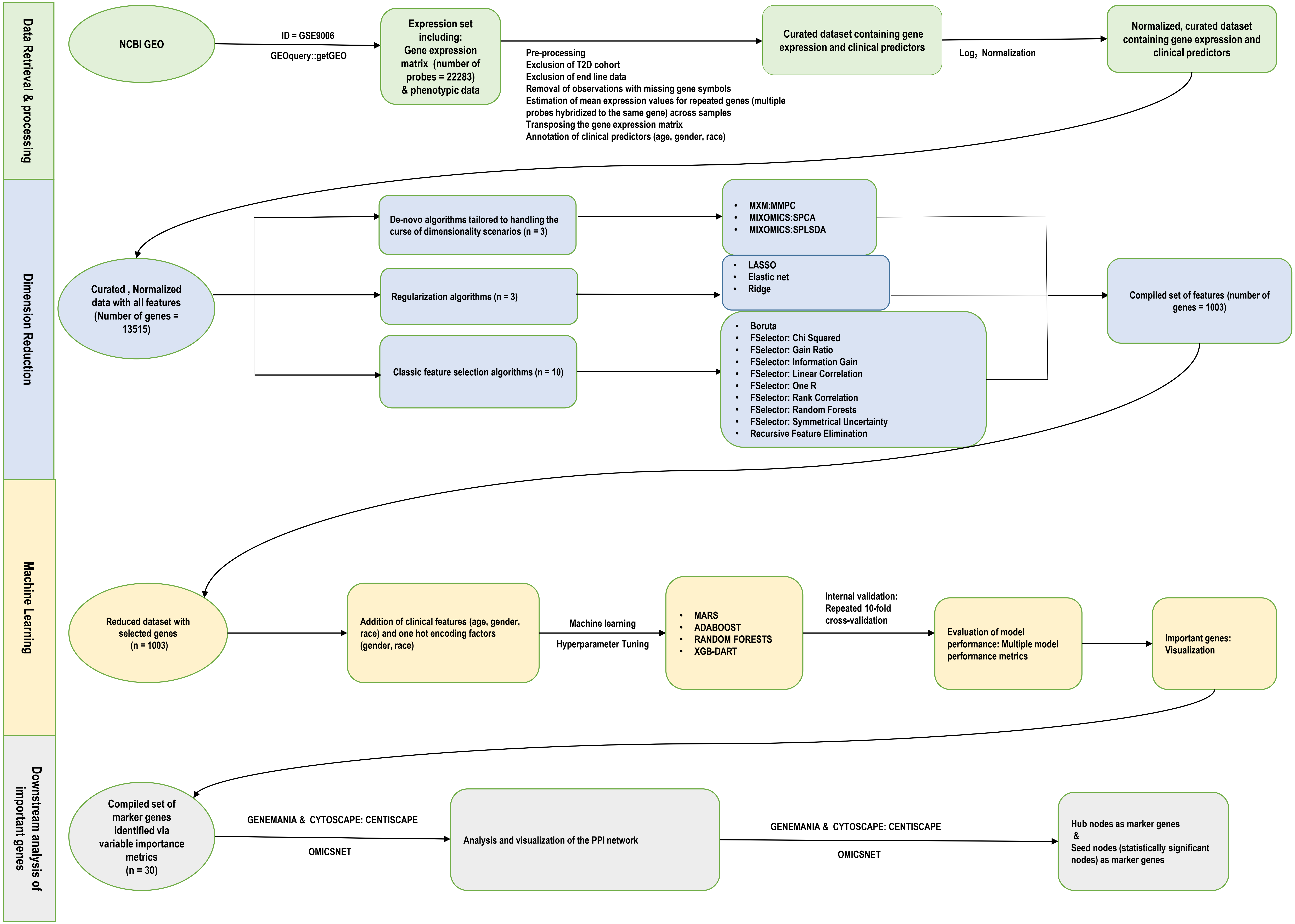
