## Supplementary Figures for "Marker genes of incident type 1 diabetes in peripheral blood mononuclear cells of children: A machine learning strategy for large-p, small-n scenarios"

**S1 Figure: Analytic workflow (Uploaded separately as “S1_Figure.pdf”)**

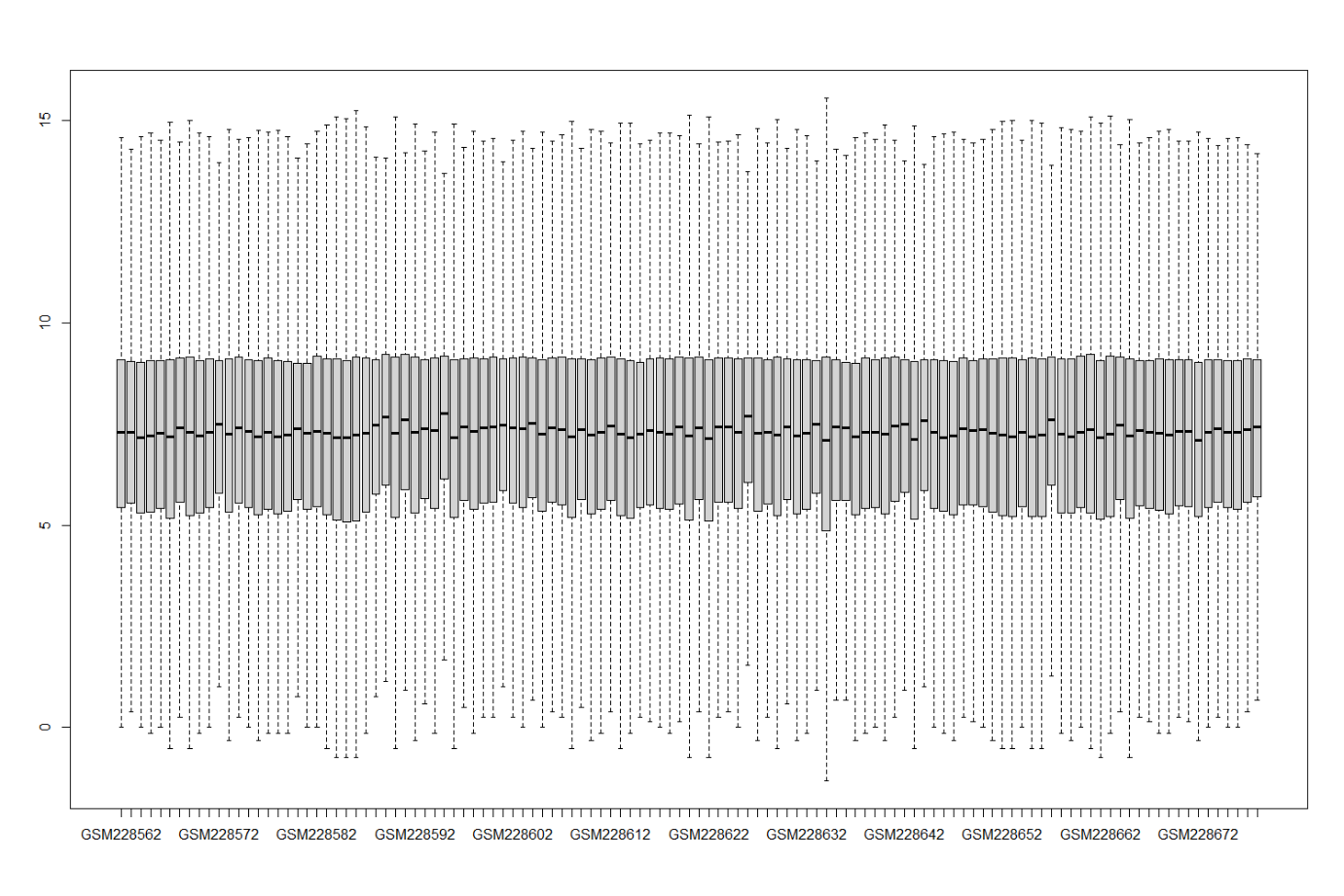

**S2 Figure: Log_2_ Normalization of gene expression matrix**

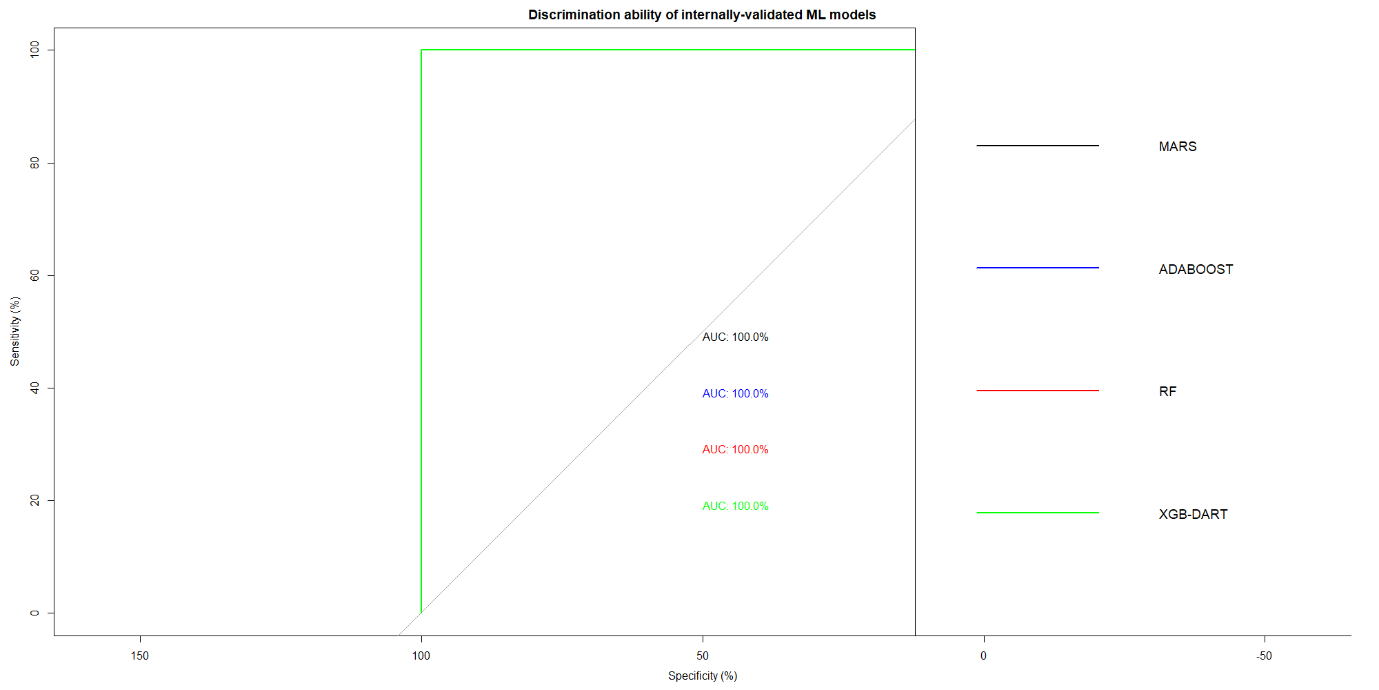

**S3 Figure: ROC curves depicting the predictive performance of internally-validated machine learning models**

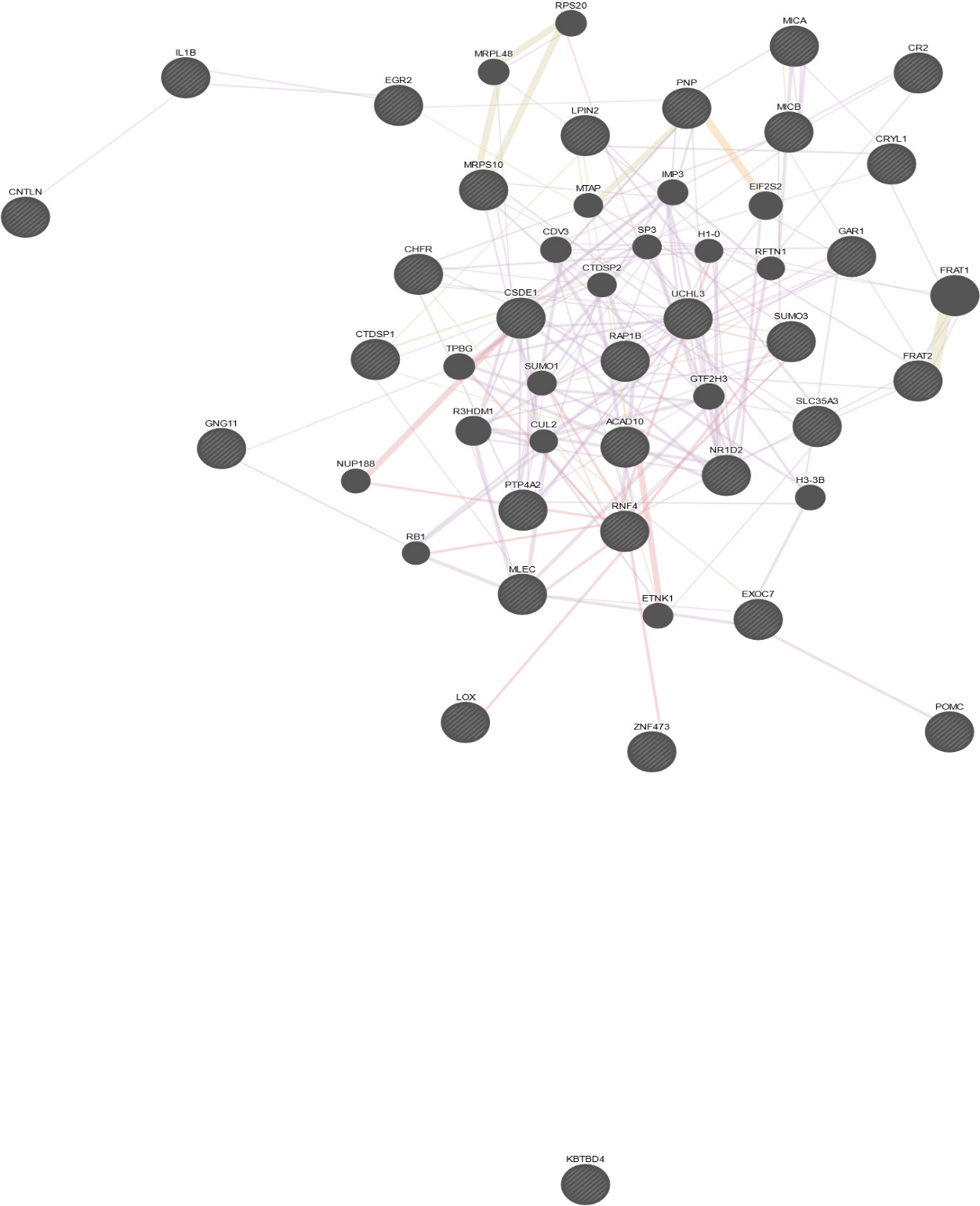

**S4 Figure: Proteins-proteins interactions network visualized in GENEMANIA**
